# Death in Venice: A Digital Reconstruction of a Large Plague Outbreak During 1630-1631

**DOI:** 10.1101/2020.03.11.20034116

**Authors:** Gianrocco Lazzari, Giovanni Colavizza, Fabio Bortoluzzi, Davide Drago, Andrea Erboso, Francesca Zugno, Frédéric Kaplan, Marcel Salathé

## Abstract

The plague, an infectious disease caused by the bacterium *Yersinia pestis*, is widely considered to be responsible for the most devastating and deadly pandemics in human history. Starting with the infamous *Black Death*, plague outbreaks are estimated to have killed around 100 million people over multiple centuries, with local mortality rates as high as 60%. However, detailed pictures of the disease dynamics of these outbreaks centuries ago remain scarce, mainly due to the lack of high-quality historical data in digital form. Here, we present an analysis of the 1630-31 plague outbreak in the city of Venice, using newly collected daily death records. We identify the presence of a two-peak pattern, for which we present two possible explanations based on computational models of disease dynamics. Systematically digitized historical records like the ones presented here promise to enrich our understanding of historical phenomena of enduring importance. This work contributes to the recently renewed interdisciplinary foray into the epidemiological and societal impact of pre-modern epidemics.

## Main text

Disease outbreaks of the plague in the past centuries have been so devastating throughout Eurasia that the term *plague* has become synonymous with a terrible disease. By killing a substantial proportion of the human population, which took multiple generations to recover, plague pandemics have had enormous impacts on the development of Eurasia. Correspondingly, historical questions, such as the role of institutions and the socioeconomic impact of plague outbreaks [1], as well as epidemiological questions, such as the causes, nature and interactions of vectors [2, 3, 4, 5], seasonality and climatic patterns [6, 7] and even the distinction between plague and the Black Death [8], are still being investigated. While previous studies have highlighted some common traits to plague epidemics [9], such as the high impact on densely-inhabited cities acting as hotspots [10, 11], the importance of human-to-human transmission [12] and the effect of the plague on different sexes [13], little is known about local outbreaks, due to the lack of detailed historical data.

We analyze high-quality data from death records created during the 1630-31 plague epidemic in Venice, whose initial investigation is limited and by now dated [14]. This epidemic was part of the so-called “Second Pandemic”, which started with the Black Death and lasted until the early 19^th^ century. Originated in northern Europe (modern France and the Rhineland) in 1623, this epidemic crossed the Alps approximately in 1629, in the case of the territories of the Republic of Venice likely carried by imperial armies on their way to Mantua. The cause of this specific outbreak in Venice has been linked to the bacterial species *Yersinia pestis* [15], and with a set of surprising results, including an uneven and unexpected impact on different cohorts by sex and age, a high parallel increase of mortality due to a synchronous smallpox epidemic and a raise in public violence [16].

Venetian death records from this period, also referred to as *necrologies*, are organized by parish and contain the systematic registration of every death among the resident population. These necrologies, edited by the parson, were established by decree since 1504 and kept in the archives of the responsible magistracy [17]. While death records were commonplace in all Christendom since the late Middle ages, and are commonly used for demography studies including on the plague [18, 1], Venetian records were particularly detailed. In the Patriarchal Archives of Venice, 54 out of more than 70 existing parishes at the time possess at least part of the registrations for the plague year (September 1630 to September 1631), while in the State Archive of Venice, the extant records for the plague year are few and scattered. Based on our assessments, these record series are overlapping and one (the former) constitutes the source for the other (the latter). We thus focus our efforts on the Patriarchal records. An example page from a necrology record is shown in Figure 1. Necrology records were kept in tiny and oblong books, with entries grouped chronologically by day. Typically, the most recurring details given for every entry were: the name, profession, sex and age of the person, the cause of death, approximate length of illness and whether a doctor attended them or not. The main dataset we use in what follows contains the number of daily deaths per parish. Data were collected following the work-flow illustrated in Figure 1; more details are given in the **SI**.

**Figure 1:**
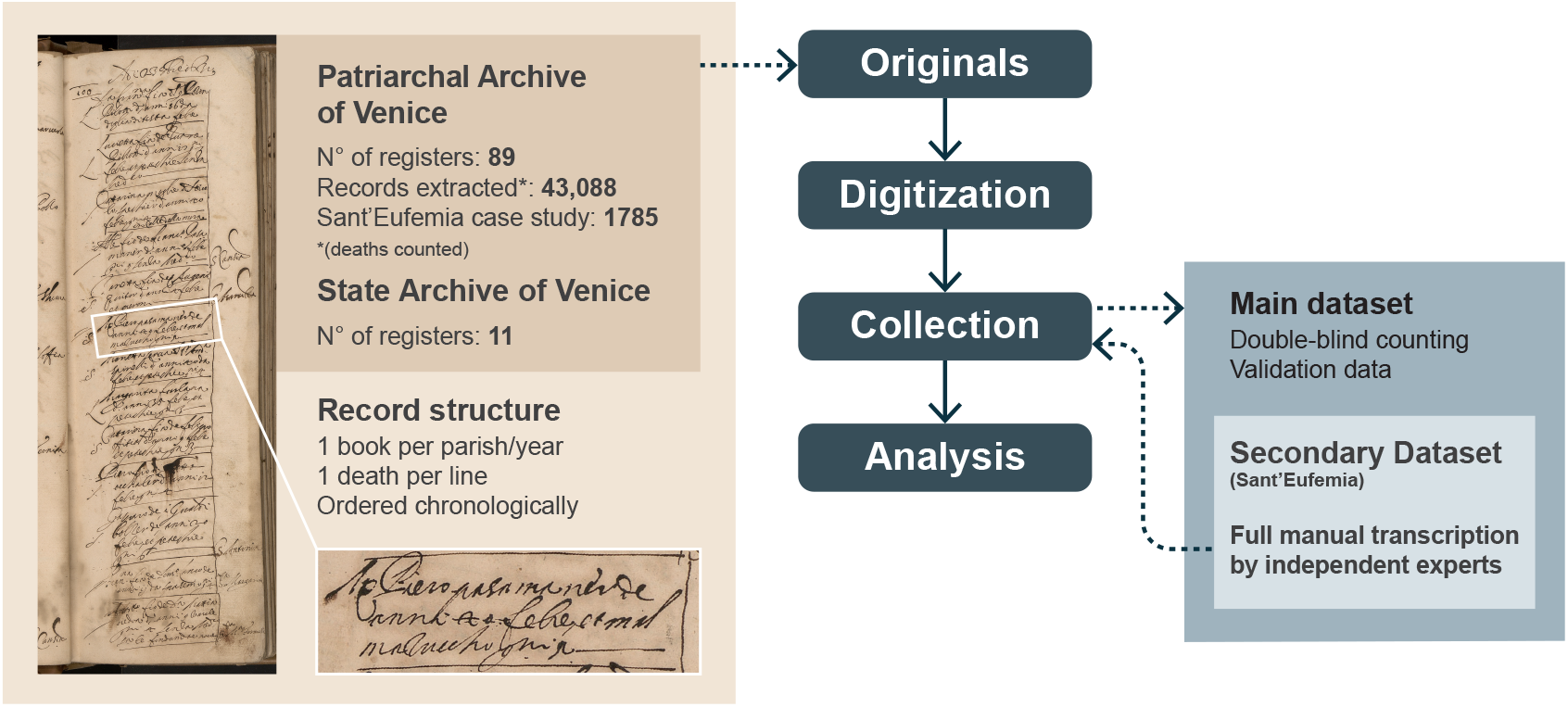
Illustration of the data collection workflow and datasets, including an example page from a death records book. The zoomed-in registration reads as follows: “*Messer Piero pasamaner de anni 40 febre et mal mazuccho giorni 5* “, which roughly translates to “Mister Piero passementerie’s weaver aged 40 fever and plague 5 days.” What is meant is that Mister Peter, a passementerie’s weaver forty of age (approximately), died of fever and plague after five days of sickness. It was the 23^rd^ of October, 1630 (as it can be read at the top of the page).

Our data aggregated over all parishes clearly shows the massive outbreak which took place between the September and December of 1630, as detailed in Figure 2a. The death counts are staggering: 20,923 deaths between September and December 1630 alone, followed by 10,430 between January and August 1631. In total, 43,088 deaths were recorded over just three years. These numbers are in line with the 35% estimated mortality in northern Italy during the same epidemic outbreak [1], and should be compared to an estimated average annual mortality between 3.7 and 2.7% (but 29.7% for newly-born infants) during the whole seventeenth century [19, 20]. We stress that not all death records survived, therefore these numbers must be taken to represent a lower bound of the actual death toll. Historical demographic sources, even though uncertain [21], report a population of 141,625 inhabitants for Venice in 1624 and of 102,243 in 1633, a reduction of 27,81% [19, 20].

**Figure 2:**
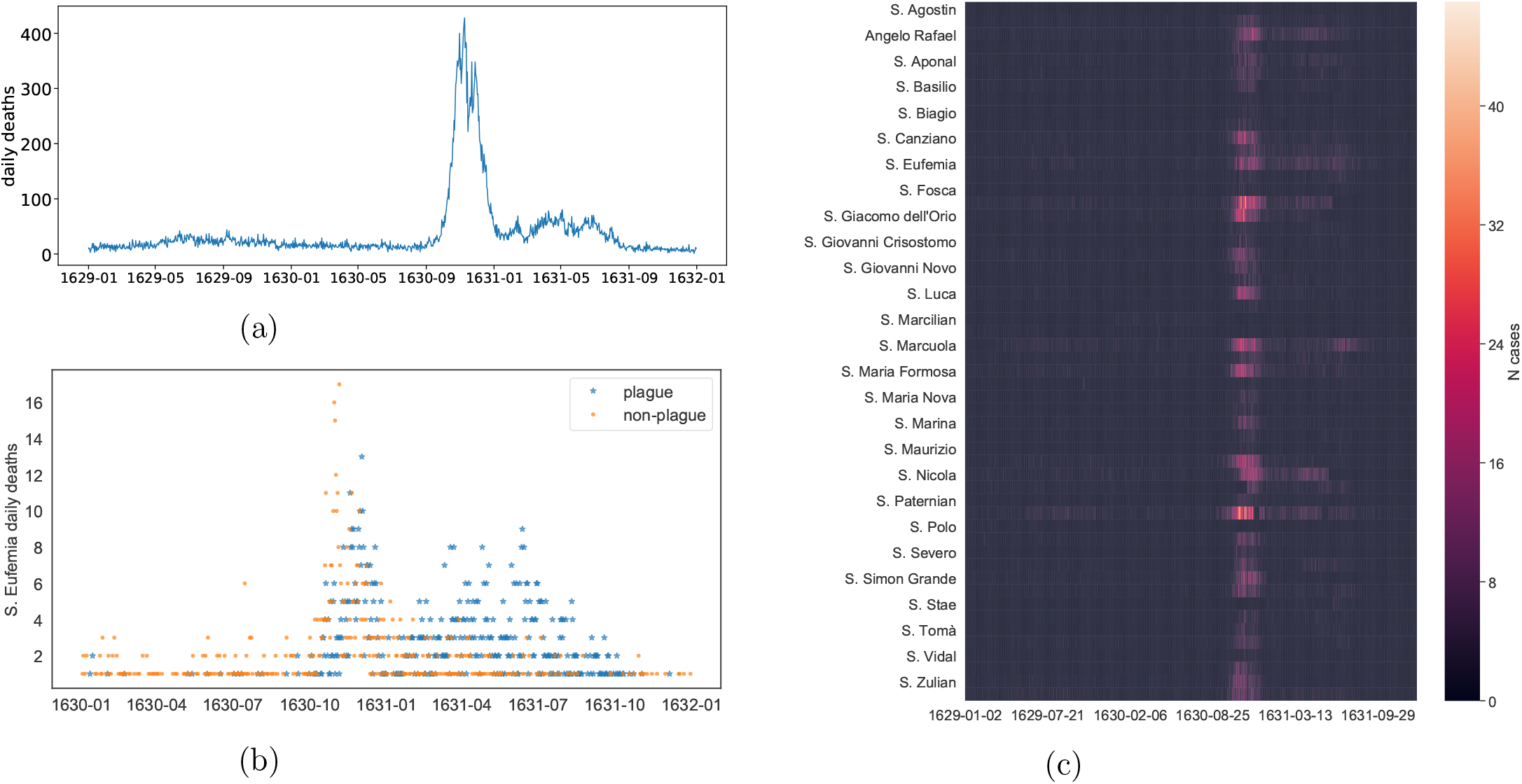
An overview of the full plague outbreak (main dataset): (a) Cumulative daily deaths for the whole recorded period (1095 days in total). A total number of 43,088 deaths were reported. One can clearly see the presence of a two-stage process, spanning until fall 1631. (b) Daily deaths recorded in the parish of *Sant’Eufemia*, almost surely due to plague (blue stars – *N*_*plague*_ = 1007) and possibly to other causes (orange circles – *N*_*not plague*_ = 778). Only days when someone died are considered. (c) A heatmap view of the dataset; for the sake of clarity, not all parishes names are plotted.

The presence of a single peak of deaths is common in plague outbreaks within densely populated regions and cities [6, 5]. Its presence in Venice indicates that the authorities’ best efforts to contain the epidemic – for example by gathering all sick people in public hospitals or in their houses [16] – simply failed. The city was too densely populated and well-connected to leave any margin for containment. In fact, as it can be seen in Figure SI1 (and especially SI1c), the outbreak in 1630 swept through the parishes practically in sync, as no discernible space correlation is present. However, while the outbreak in 1630 is known, the subsequent 1631 long tail of high mortality has not been described in the literature before.

In order to gain a better understanding of the disease dynamics, we investigated another dataset taken from the records of a specific parish: *Sant’Eufemia*. This was a populous parish, with a significant amount of deaths in the 1631 tail and whose necrology records are well-preserved in their entirety. We transcribed all the information available in its necrologies, i.e. the name, sex and age at death of each person, together with the cause of death and the length of sickness. This transcription includes 1785 deaths registered between January 1630 and December 1631. The identification of deaths due to plague appears to be deceptively simple, as they were usually registered as fatalities caused by suspicious illness (“*mal sospetto*”), or with visible buboes. Nevertheless, previous studies have taken a more inclusive approach, considering also deaths not clearly caused by other factors as due to plague [16]. We take the more conservative approach in what follows – see Tables SI1, SI2 and SI3 for details on which causes of death were considered to be plague.

The statistics of the causes of death give us a first insight. In Figure SI2a we show the distribution of deaths grouped by cause and (conservatively) classified as related to the plague or not. One can see how the two distributions are skewed, meaning that a small fraction of causes (5%) contributes to a large fraction of deaths (63%). However, while the number of deaths clearly due to plague (*N*_*plague*_ = 1007) and possibly non-plague are similar (*N*_*not_plague*_ = 778), only 56 out of 156 causes could be clearly attributed to plague, leaving more vagueness around the non-plague causes (in Figure SI2b the causes with more than 50 deaths are listed). This seems to suggest that our plague-death counts likely constitute a lower bound of the total number of deaths directly linked to plague, which we cannot further refine from the records. In Figure 2b we show the time-series of deaths belonging to the *Sant’Eufemia* parish, distinguishing between those caused by the plague and the ones possibly due to other causes. Surprisingly, the first peak of the epidemic begins with few references to the common symptoms of the plague (October to November), when the records point instead to more generic and common illnesses, such as fever or spasms [17]. Only afterwards the records start to extensively mention the plague as the cause of death, well into the Fall of 1631. This might indicate an initial reticence to acknowledge the epidemic outbreak, as well as a subsequent possible overemphasis of it. This reticence might be caused by the public authorities’ practice to quarantine the whole household in their house when someone from it died of plague. It might also be due to a surveillance issue generating a bias in the records: while many deaths were occurring, medical examination was no longer taking place and the registrations of the causes of death were not happening regularly, but instead in batches, leading to approximations. Furthermore, several people were moved to quarantine areas (*lazzaretti*) and died there, while their registration happened subsequently, possibly by reporting generic causes of death. It is thus likely that these deaths are also in large part attributable to plague. However, other explanations are also possible, such as a known epidemic of smallpox co-occurring during the main peak [16]. Despite these limitations and open questions, *Sant’Eufemia*’s causes of death confirm the duration of the epidemic well into the autumn of 1631.

We further verify that deaths by plague were not significantly affected by sex, under the reasonable assumption that sexes were equally distributed in the population of Venice at the time [20]. Indeed, the male to female deaths ratio was close to one (*N*_*male*_*/N*_*female*_ = 865*/*917 ∼ 0.94), a result confirmed by the majority of the literature [1, 12, 22, 23, 24, 25], with few exceptions [16, 13]. Furthermore, the distribution of illness duration and of age at death did not significantly change with sex (see Figure SI3a and SI3b respectively). Assessing the effect of the plague on age is challenging, as assumptions on the age distribution of population at that time are quite difficult to make and historical statistics are hard to find. Furthermore, the literature on the effect of the plague on different age cohorts is still ambiguous. Nevertheless, our data are in line with previous studies [26, 27, 18, 28, 1] indicating that the plague had higher relative impact among age cohorts of typically low mortality, in particular adolescents and adults between 14 and 44 years of age, as shown in Figure SI3c and Figure SI3d.

Figure 2c shows the heatmap of reported cases, for each of the parishes of Venice, for the entire time window (*N*_*tot_deaths*_ = 43088). One can see that while the main outbreak occurring in the last four months of 1630 shows good synchronization across all parishes, the second, smaller outbreak occurring until fall 1631 seems to have peaked at rather different time points within each parish, between February and July 1631. We therefore investigate whether space patterns are present, especially in the 1631 outbreaks (*N*_*deaths_tail*_ = 10363). In order to assess the presence of spatial patterns, we simply plot the pairwise correlation among cases for all couples of parishes, against the distance between parishes (Figure SI4). The resulting scatter plots show no spatial patterns. Nevertheless, the secondary outbreak in 1631 does not seem to have peaked as homogeneously as the first large outbreak in 1630 (Figure 2c). We hence performed a clustering analysis to highlight possible groups of rather synchronous parishes (Figure 3). The analysis on the main 1630 outbreak (Figure 3a) appears instead to be in sync across parishes.

**Figure 3:**
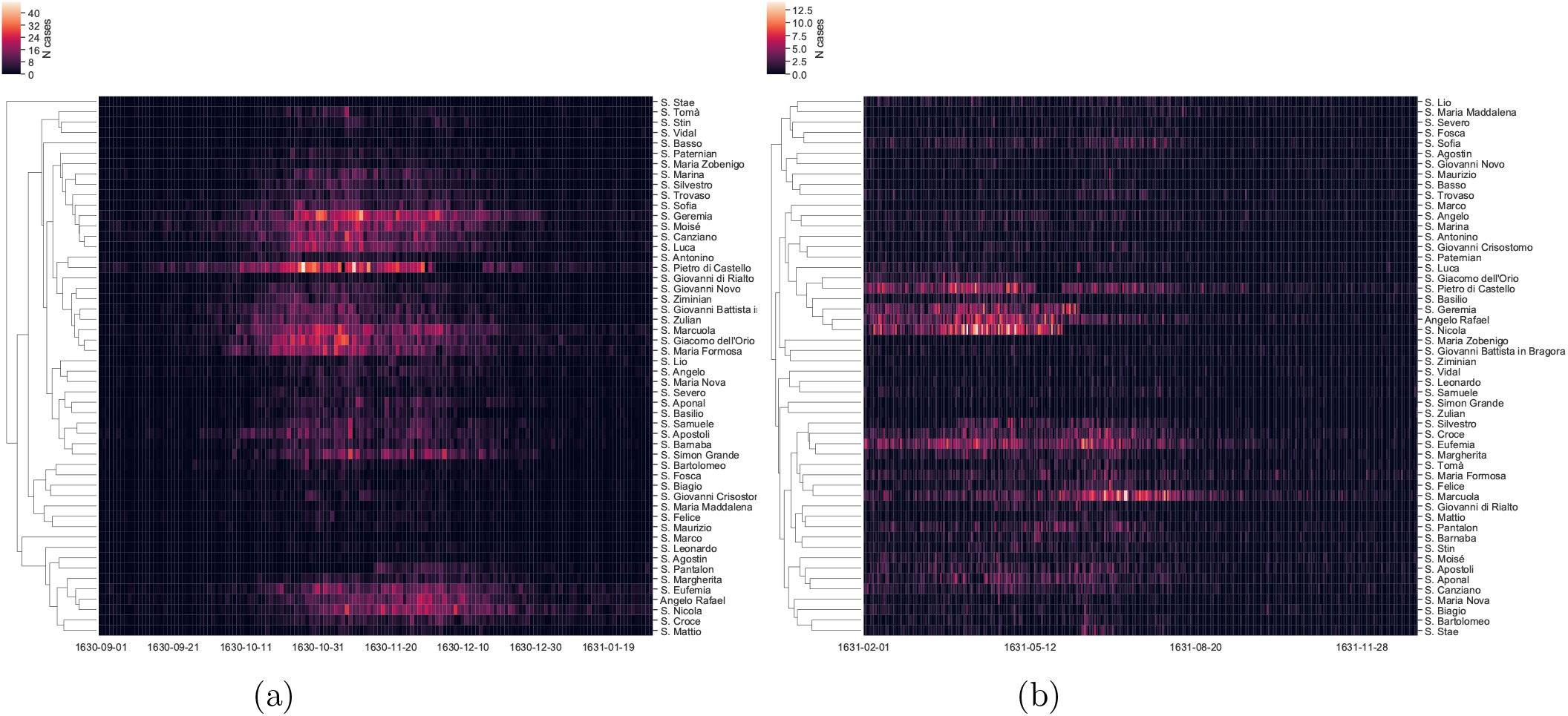
Hierarchical clustering of parishes zoomed on the main late-1630 peak (a) and on the 1631 outbreaks (b).

The clustering on the 1631 outbreaks (Figure 3b) shows clusters of parishes with more spread-out peaks, across the first half of 1631, with tails reaching the fall of the same year. The main cluster is the one led by the three populous parishes of *S. Geremia, Angelo Rafael* and *S. Nicola*, with peaks between March and May 1631 (central part of Figure 3b). Another cluster is the one led by the *S. Eufemia* and *S. Marcuola* parishes (bottom part of Figure 3b), a more heterogeneous group, with peaks occurring mostly in June/July 1631.

Even though these clusters seems to be well separated in time, there is no clear evidence of a specific process or event in the history of the city that might have driven this spatial distribution of localized epidemics in different parishes during 1631. We therefore assess epidemiological models on data aggregated over all parishes. The plague is generally modeled as a zoonosis, in which the transition from an epizootic (typically, in rodents) to a human epidemic is mediated by animal fleas, the vector carrying *Yersinia Pestis* [30, 29]. *From here on, we refer to this model as the Rats-Fleas-Humans (RFH) model. At the same time, other studies suggest that these models are not always preferable to explain the outbreaks dynamics, especially due to the ‘efficacy and speed’ of some historical plague* outbreaks [1], if compared to the typical dynamics of RFH models. We first confirm that neither a deterministic RFH nor a deterministic Susceptible-Infected-Removed (SIR) model can explain the presence of the 1631 secondary outbreaks (see Figure SI5b). We then investigate the transmission nature of the Venice plague, by considering separately the main 1630 outbreak and the one in 1631. In both cases, we find that the RFH model did not perform much better than a simple SIR model, as shown in Figure 4a (main 1630 outbreak), and Figure SI5c (1631 outbreaks). We therefore implement a time-dependent SIR and find that it can better explain the dynamics over the entire time window (Figure 4b), with an increase in the basic reproduction number that could indicate a change in the transmission mechanism of pathogen (for clarity, fitted parameters are reported in Figure SI5d). In particular this might suggest a transition from bubonic to pneumonic plague, a shift already hypothesized for other historical plague epidemics [31]. However, a change in the effective transmission rate might also be due to people’s behavioral response to the outbreak. In order to investigate the fitness of such hypothesis, we implement a stochastic delayed behavioral SIR (details can be found in the `Methods`). In Figure 4c we show one example of such model’s stochastic realizations, which presents both a main peak and a long tail dynamics. This shows that a change in pathogen’s transmission route is not necessarily required in order for the epidemic to show a non-trivial temporal pattern, such as the one present in our data. For the sake of completeness we also check whether a deterministic delayed behavioral SIR would fit our data. In Figure SI6b, we show that it cannot actually reproduce the 1631 tail, in spite of a good fit of the first part of the 1630 outbreak.

**Figure 4:**
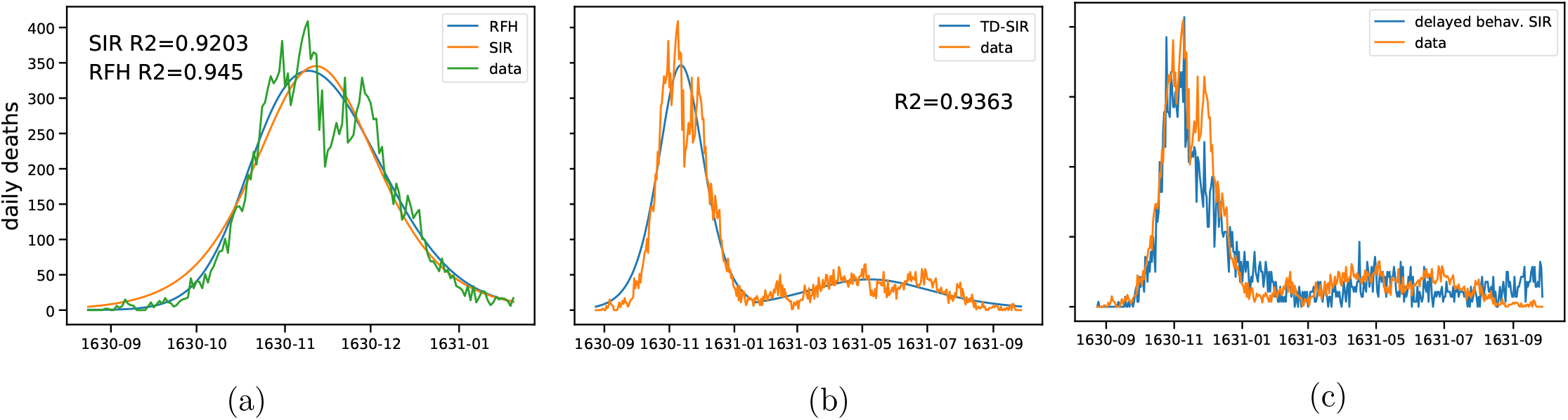
(a) Best fit comparison of a simple SIR model against the model from [29] on the main outbreak peak (150 days time window). (b) Best fit of an explicit time-dependent SIR; parameters are shown in Figure SI5d. (c) Example realization of a stochastic delayed behavioral SIR; the evolution of transmission rate *β*(I) is shown in Figure SI5e.

Although a change in diffusion parameters seems to provide a reasonable explanation of the two-peak structure, we investigate the possibility of having two-peak outbreaks similar to the observed one, as a result of the stochastic nature of the disease spread combined with structural properties of the host network. It is indeed known that the community structure of a network can strongly impact epidemic dynamics [32]. We therefore perform a series of stochastic simulations of a simple SIR process on top of a small-word graph, a network model which is likely to resemble the modular structure of social contacts [33] (further details on the simulations are given in the `Methods`). We find that few simulated epidemics do resemble the data, as shown in Figure SI6a. However, as this happens in only about 0.1% of the simulations, such alternative interpretation of the 1631 tail based on pure stochastic effects and network structure, although reasonable, remains very unlikely.

In summary, we find a novel epidemic pattern of two peaks in the 1630-31 plague outbreak in Venice. The first peak in 1630 was very high, and the outbreak highly synchronized among all parishes; the second peak in 1631 shows temporal variability, and was much less pronounced in strength. Most previous recorded cases show a single main peak [6, 29, 5] of varying duration [18, 12], with possible cyclical recurrence [6]. Relying on fine-grained daily death records [1], we are able to confirm that the plague spanned both the main peak and the long tail, over a period of more than a year and caused the death of approximately 30% of the city’s population.

Providing an interpretation of the two-stage process remains challenging with the evidence at our disposal. Firstly, not all deaths could be clearly attributed to the plague during the early weeks of the main peak. Generic causes of death such as fever and spasms might indicate plague deaths as well as deaths due to other causes. A first hypothesis is therefore that the same plague epidemic went on for more than a year, while being aggravated by other concomitant causes during the main peak. An alternative hypothesis is that two distinct plague epidemics took place instead, one during the main peak and another during the long tail. Previous studies suggest the possibility of a transition from a mainly bubonic to a mainly pneumonic plague, for example. Furthermore, we show that it is also possible that such temporal pattern could be generated by the adaptation of hosts’ behavior to the increase of number of infected, effectively decreasing the transmission rate, as the outbreak advances. Lastly, social factors such as the timing and effectiveness of public containment policies could have played a role.

Further investigations will be needed in order to fully qualify the Venetian 1630-31 plague outbreak, as well as the Second Pandemic overall. Indeed, as we have shown, historical records contain information which has so far been relied upon only to study few episodes but, when digitized and made available at scale and systematically, can help cast new light on these long-lasting research issues. For an understanding of detailed local dynamics, but also of global patterns of disease spread, modern human data and animal research can now be complemented with digital data collection driven by the digital and medical humanities.

## Data Availability

Data will be shared in the following months, together with code for data analysis and simulations

## Author contributions statement

G.L. and G.C. performed the analysis. D.D., F.Z., F.B., A.E. and G.C. performed data collection. G.L., G.C., M.S. and F.K. wrote the paper. M.S. and F.K. supervised the study.

## Acknowledgements

We would like to thank the support of the Patriarchal Archive of Venice and the State Archive of Venice during data collection. We thank Paolo de los Rios and Giulio Rossetti for useful discussions on diffusion processes on networks, and gratefully acknowledge the help of Laurent Bolli in designing Figure 1.

## Methods

### Data collection

The main dataset we consider consists of the daily number of deaths per parish, from January 1629 to December 1631. We have first proceeded with a full double-blind counting, then compared the two series, checking and correcting all discrepancies. Secondly, two different co-authors have counted again all deaths from a sample of 20 parishes out of 70 (8 and 12 each), to further assess our main dataset, with the following results:

- 1629: 22 errors over 2395 assessed registrations (0,91%).
- 1630: 60 over 8989 (0,66%).
- 1631: 16 over 3730 (0,42%).

Confirming that the main dataset was already of high quality. Eventually, all remaining errors were checked again and corrected in the final dataset, which we analyze in this contribution.

We note that the parson of every parish was supposed in principle to a) get a medical inspection of every dead body to rule out contagious causes, b) report all deaths every morning to the magistrate called *Provveditori alla Sanità*, c) get burial licenses from this magistracy before inhumation. Steps a and c usually were not taking place during the months of peak mortality at the end of the year 1630. It is important to clarify that our death records include deaths which occurred in the main care institutions in Venice: the four *Ospedali Grandi* (main hospitals), as well as minor ones, with respect to residents in the available parishes. They also include all deaths occurred at the *lazzaretti* : temporary locations setup for quarantine or inhumation of persons affected by the plague. They do not include foreigners. We finally note that the parish of *S. Nicola* is to be identified with *San Nicola dei Mendicoli*.

### Data analysis and modeling

All data analysis and modeling are done in Python. For the general data cleaning we use the `pandas` package. The distance between two parishes is defined as the geodetic distance between the centers of the corresponding polygons, defining the jurisdiction of the same parishes. The `geodesic` function from the `geopy.distance` module is used for this task.

All dendrograms (Figure 3) are plotted using the `seaborn.clustermap` package. In particular, we use the metric `correlation`^1^ and the method `complete` to build the linkage matrix, needed to compute the clusters. The compartmental epidemic models are integrated using the `odeint` function from the `scipy.integrate` module. The parameters estimations are then obtained using the `curve fit` and `differential evolution` function from the `scipy.optimize` package [34]. In order to account for false positives, we estimate a baseline of deaths very likely to be unrelated to the plague outbreak, by fitting a sinusoidal signal from the beginning of the recordings, until the end of August, as shown in Figure SI5a. In the time-dependent SIR model we assumed a simple step function dependence for both *β*(*t*) and *γ*(*t*), leading to a total of five fitted parameters: *β*_1_, *β*_2_, *γ*_1_, *γ*_2_ and the transition time *τ* (see again fig SI5d).

Stochastic simulations in Figure 4c and SI6a were done using the `ndlib` package [35], on graphs generated with the `networkx`package [36].

The delayed behavioral SIR model (Figure 4c) was defined using the following expression for the transmission rate *β*(*t*) = *β*_0_*e*^−*I*(*t*−*τ*)*/I*^*, where *β*_0_, *τ* and *I*\* were fitted parameters, together with the usual (constant) death rate *γ* and initial number of infected *I*_0_ (*β*_0_ = 0.06429, *I*\* = 72, *τ* = 32, *γ* = 0.02859, *I*_0_ = 3). For its stochastic implementation we used a Erdos-Renyi graph, with an edge creation probability *p* = 4*/N*_*nodes*_(*N*_*nodes*_ = 20000).

## Data availability

All code and data needed to reproduce plots and analysis presented in the manuscript will be made available in a dedicated GitHub repository before publication.

## Supplementary Information

**Supplementary Figure SI1:**
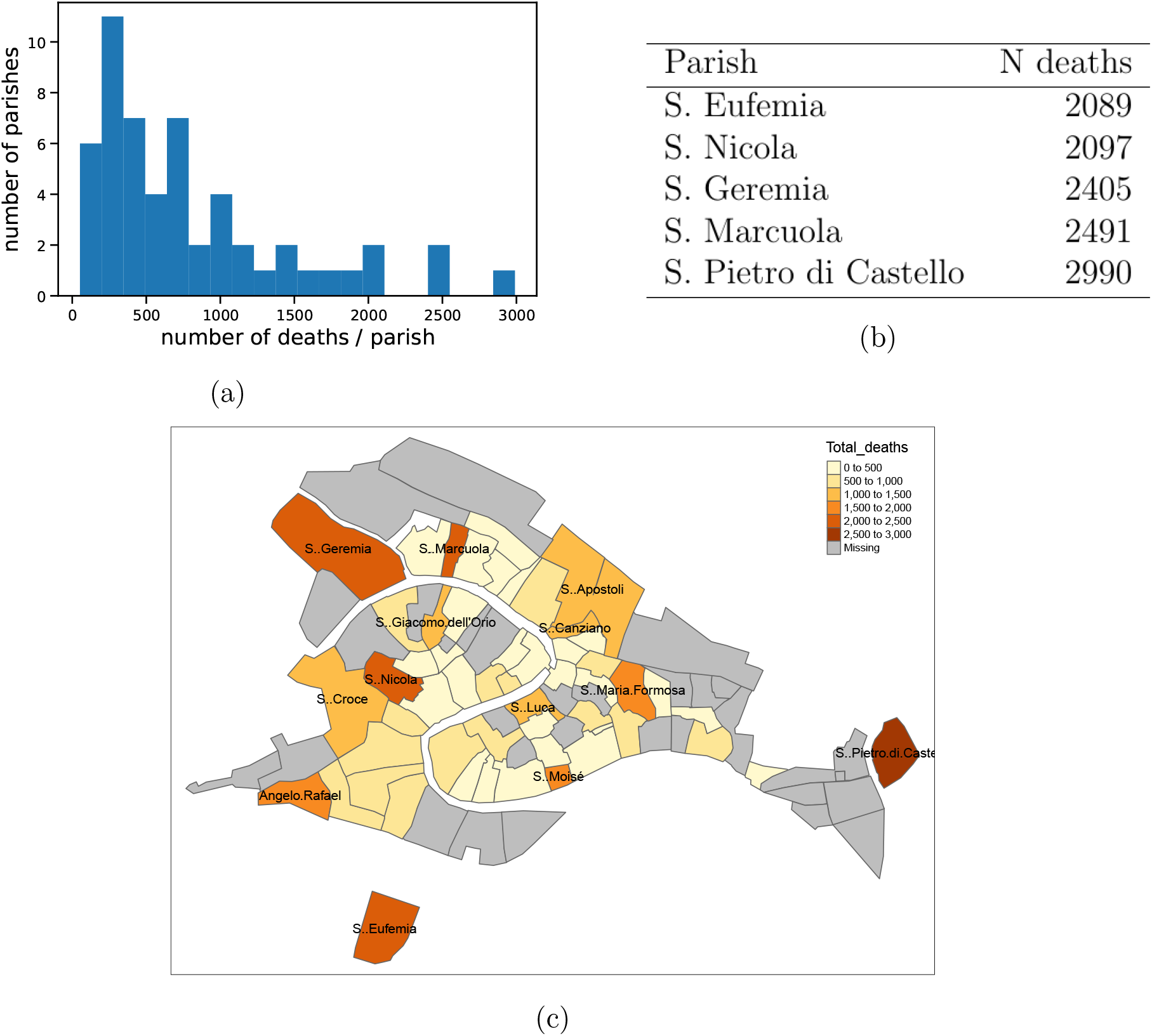
Distribution of number of deaths by parish. (a) One can clearly see the skewed distribution, with the top 24% of parishes accounting for about 55% of the total deaths. (b) For the sake of clarity, only parishes with more than 2000 deaths are listed. (c) Map of Venice parishes, color-coded by total recorded deaths, summed over the entire time-window. For clarity, only the names of parishes with more than 1000 total deaths are shown.

**Supplementary Figure SI2:**
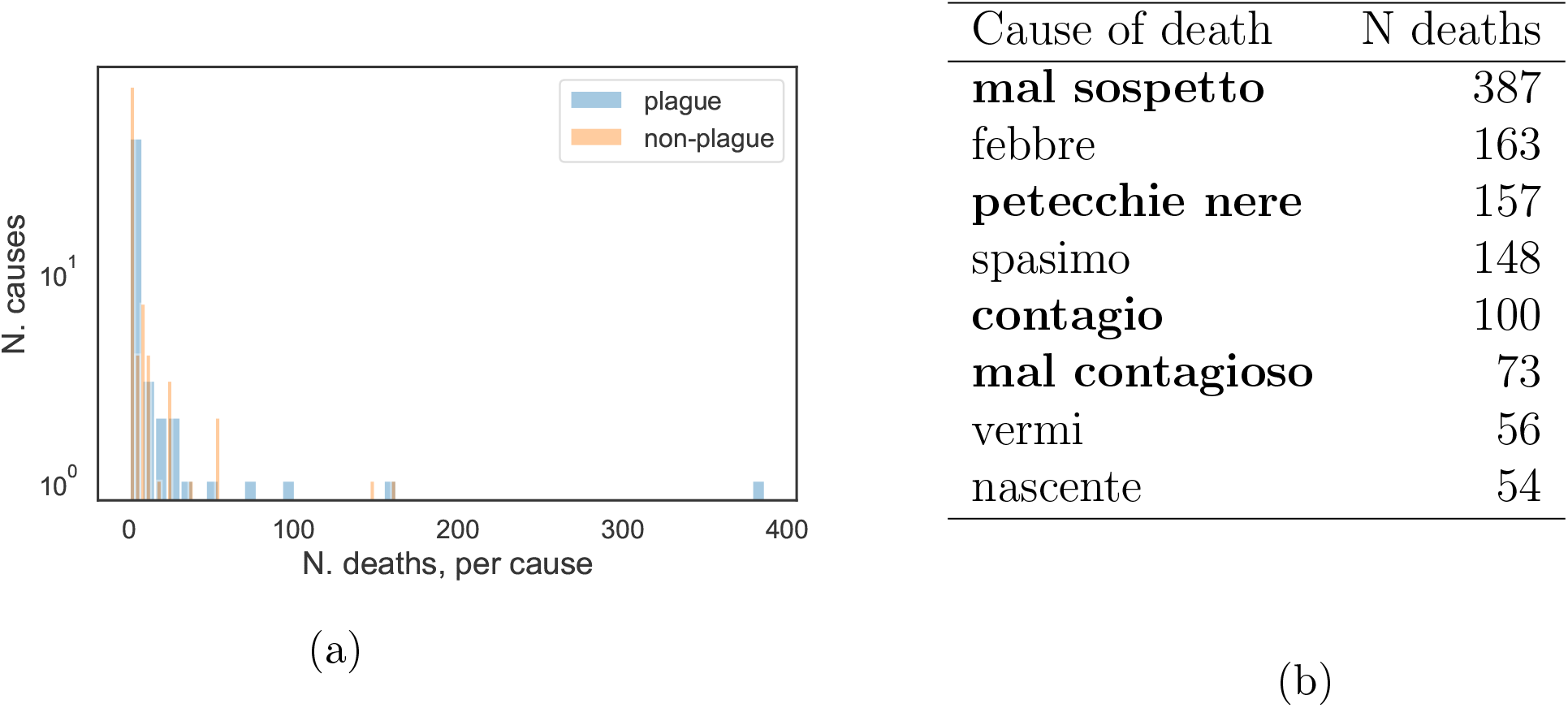
Distribution of number of deaths by cause, for the parish of *Sant’Eufemia*. In the records, 156 unique causes of death are found, of which 56 were attributed to plague. One can clearly see the skewed distribution, with the top 5% of causes accounting for about 63% of the overall deaths (a). For the sake of clarity, only causes with more than 50 deaths are listed; in bold the ones attributed to plague (b).

**Supplementary Figure SI3:**
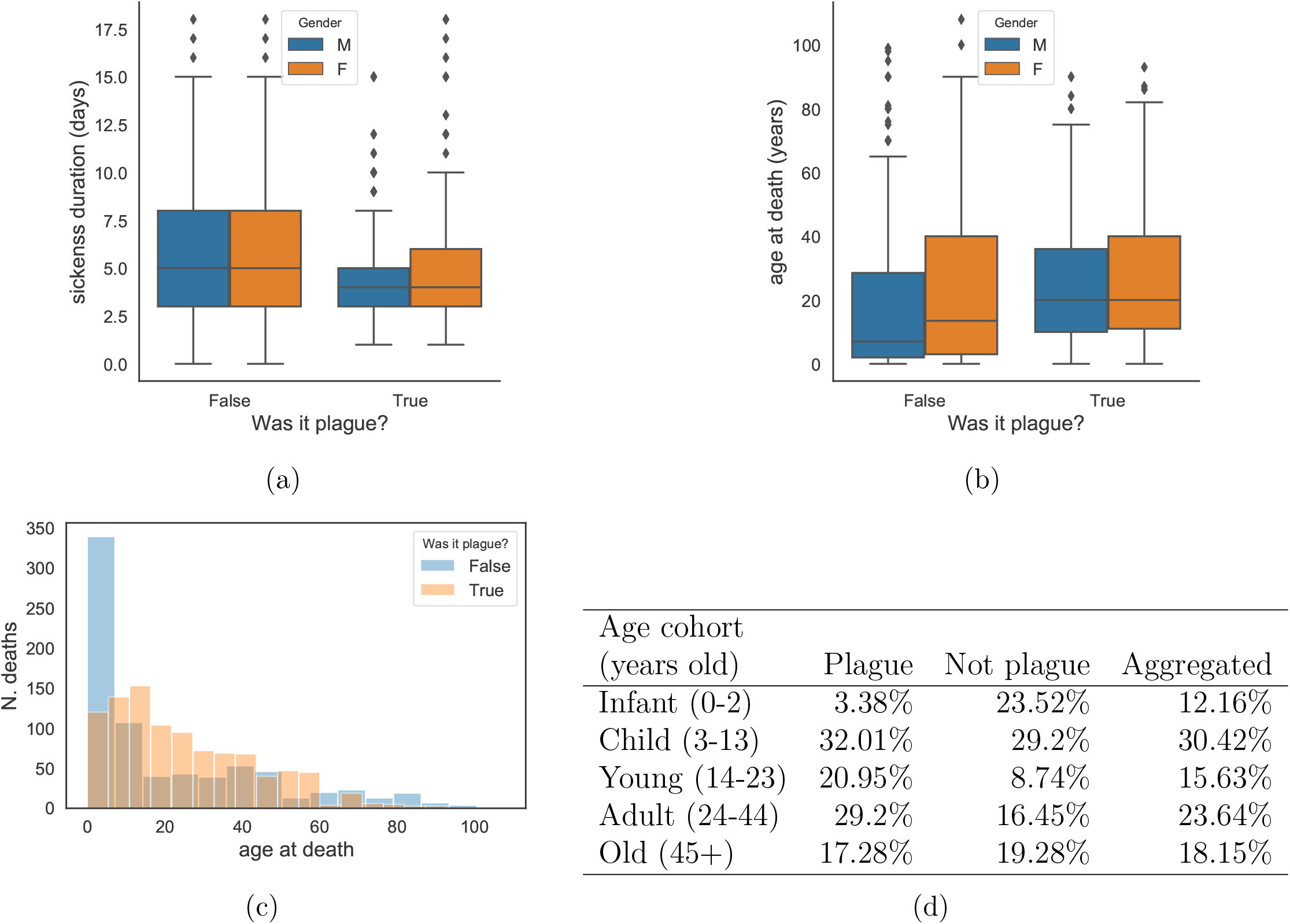
Demographic statistics for the parish of *Sant’Eufemia*. (a) Distribution of sickness duration, by sex and cause of death for *S. Eufemia* death records – for sake of clarity the boxplots include only cases with a sickness spanning less than 20 days (this still covers about 88% of the total sickness duration distribution). (b,c) Distributions of age at death, divided by cause of death. No significant age difference emerges due to sex (*p >* 0.001 on two samples KS test, for both causes of death) (b), while a significant one appears between the plague VS non-plague deaths, aggregated over sex (*p* < 10^−20^ on two samples KS test) (c). (d) Table with the same numbers of deaths, divided into age groups. Note that an infant mortality at birth between 20 and 30% was common at the time [20].

**Supplementary Figure SI4:**
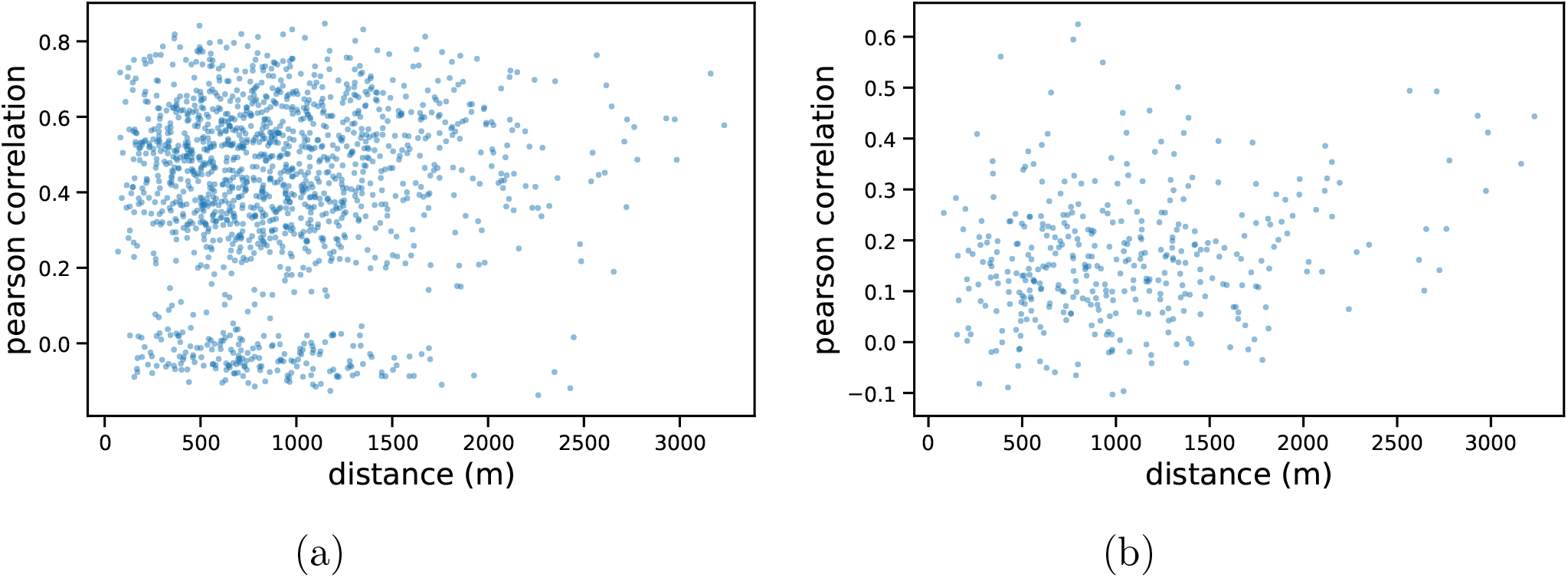
Pairwise Pearson correlation between cases time-series of each couple of parishes, as function of distance between the two parishes. The same scatter plot for all parishes (*N*_*parishes*_ = 54) for the entire time-windows (a) and for the largest parishes, for the 1631 outbreaks only (b). The largest parishes are defined as those reporting more than 500 deaths (*N*_*parishes*_ = 28).

**Supplementary Figure SI5:**
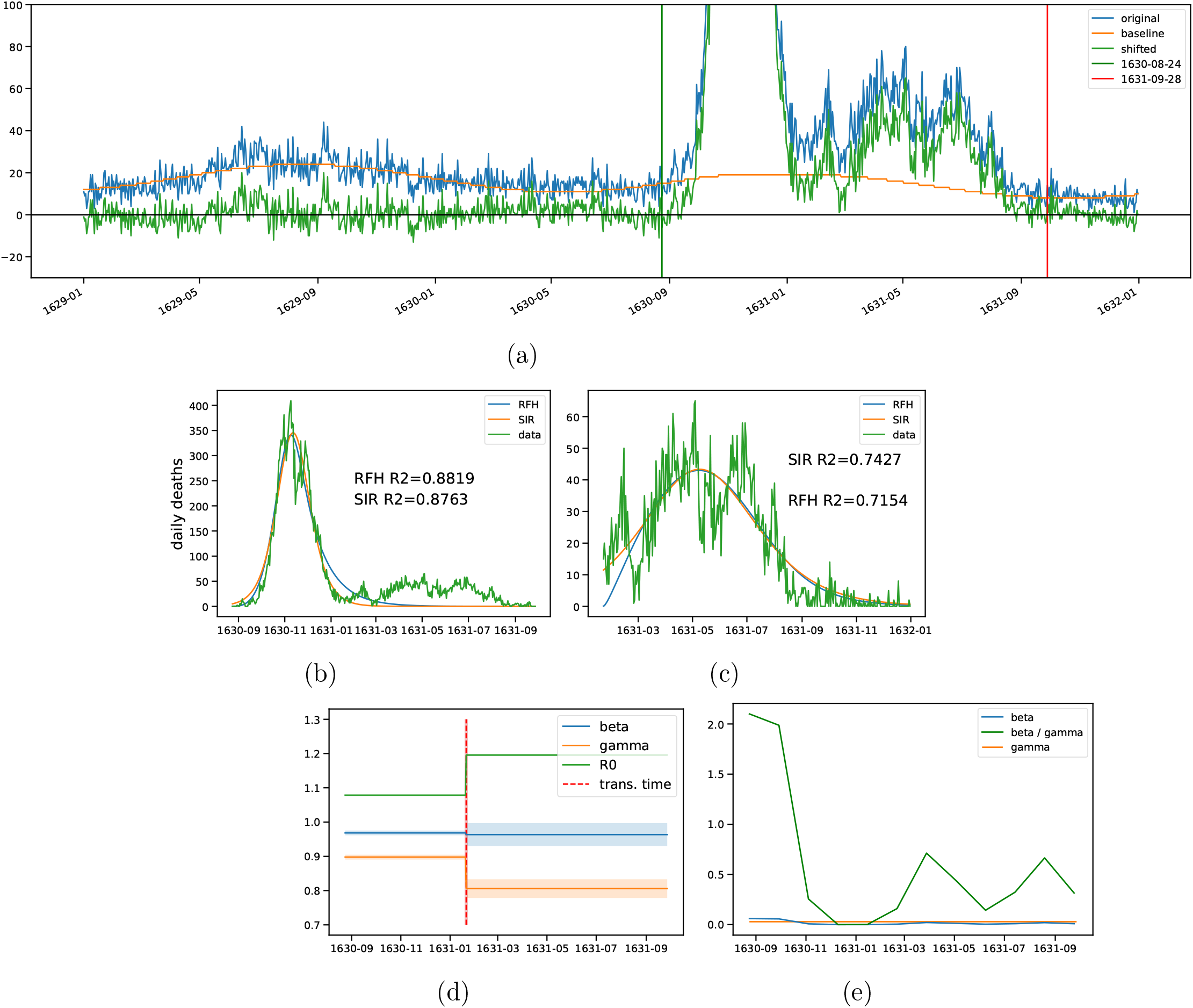
(a) Estimation of baseline cases due to other causes: a sinus function (orange line) is fitted from the beginning of the data until the beginning of the fit (green vertical line) to estimate the mortality rate, which is then applied to the original data (blue) to get the data used for the fit (green). (b-c) Comparison between a simple SIR and the more complex RFH model, in the 400 days window (b) and zoomed on the second part of the epidemic (c). (d) Fitted parameters for the time-dependent SIR model, as in Figure 4b. (e) Evolution of fitted *β*(*I*) and *β*(*I*)*/γ* for the delayed behavioral SIR shown in Figure 4c.

**Supplementary Figure SI6:**
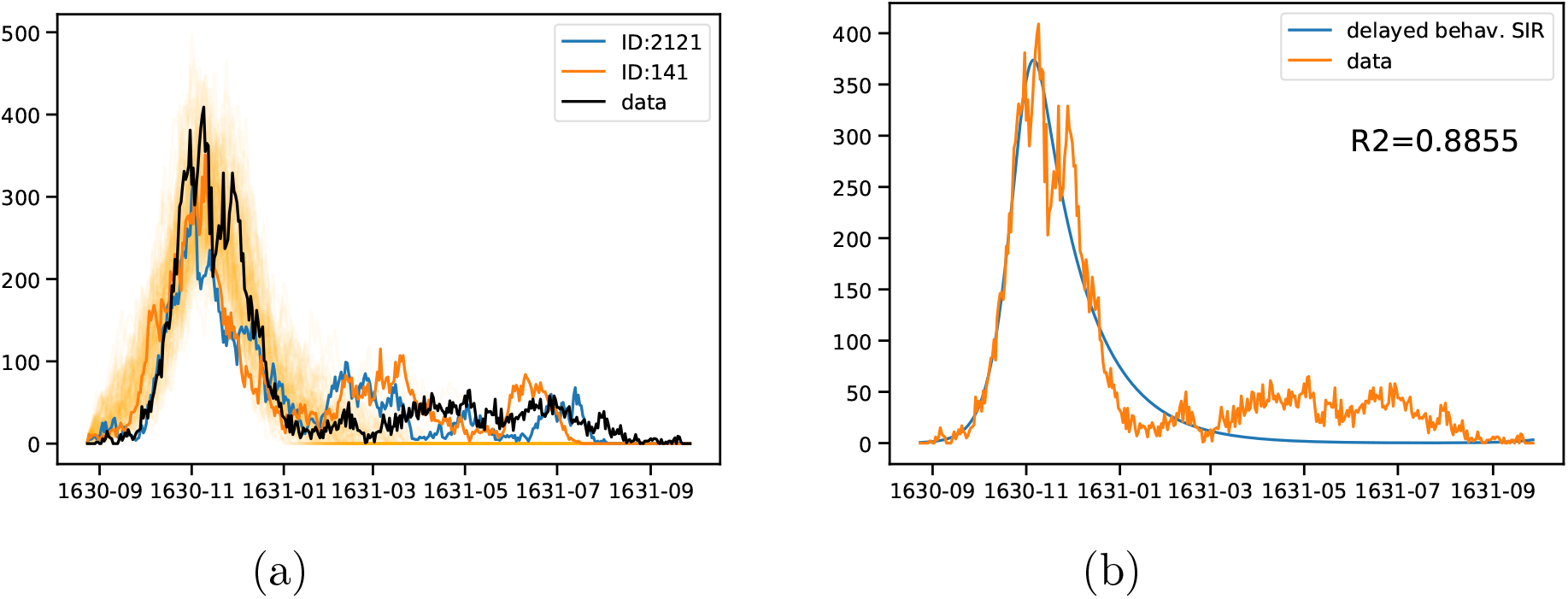
(a) Selected stochastic simulations of a simple SIR model on top of a small-word network. Two particular epidemics are highlighted, in order to show the possibility of having a large peak followed by a long tail, as present in the data. In shaded orange we only show, for sake of clarity, the simulated epidemics with lowest deviation from the data (*RMSE* < 50 −*N* = 93) (b) Best fit of deterministic behavioral delayed SIR. Although the model can fit very well the first part of the epidemic, it does not show a secondary outbreak, in 1631.

**Supplementary Table SI1:**
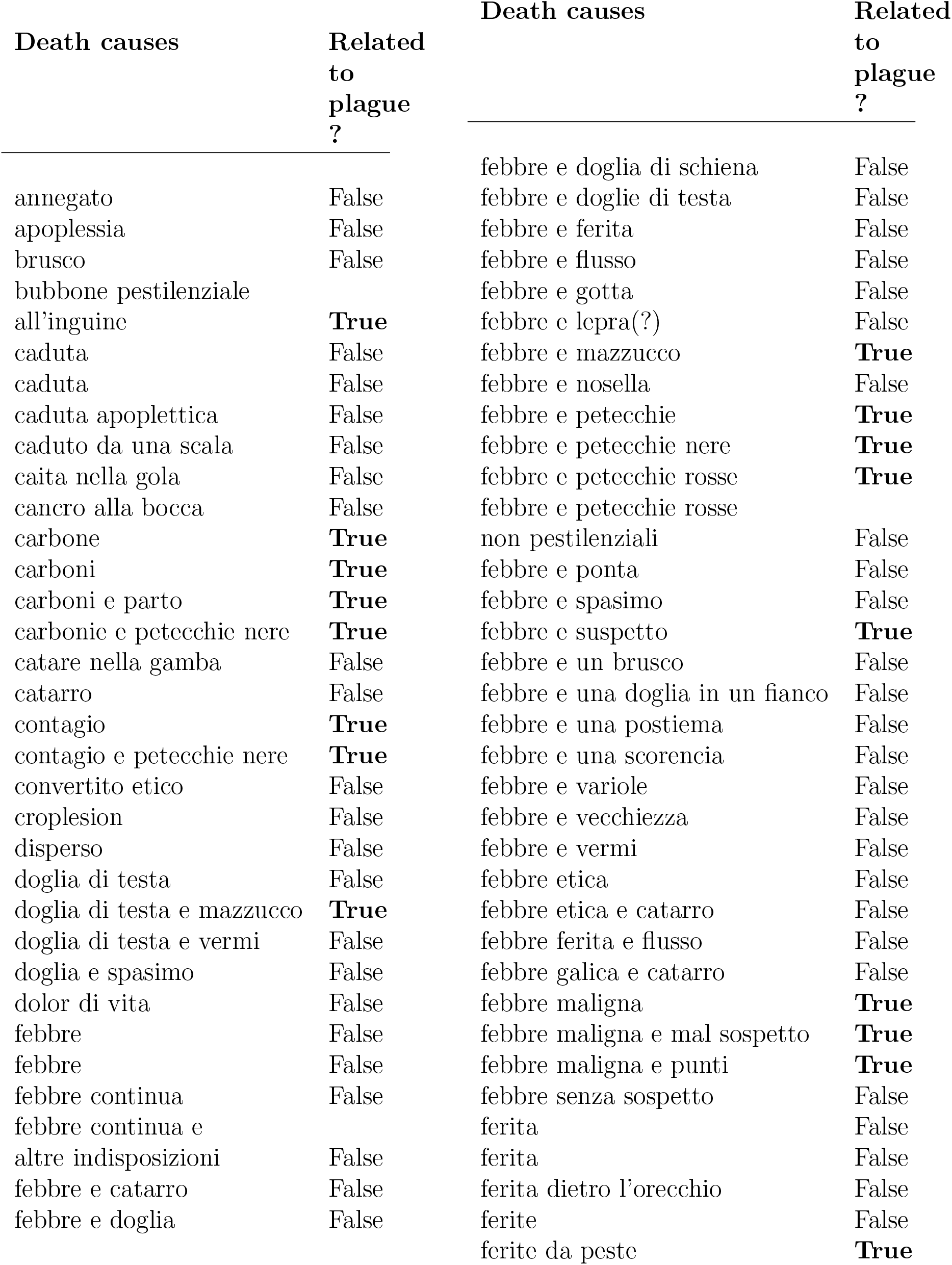
Manual classification of all 156 death causes as reported in necrologies, as associated to plague or not – part 1.

**Supplementary Table SI2:**
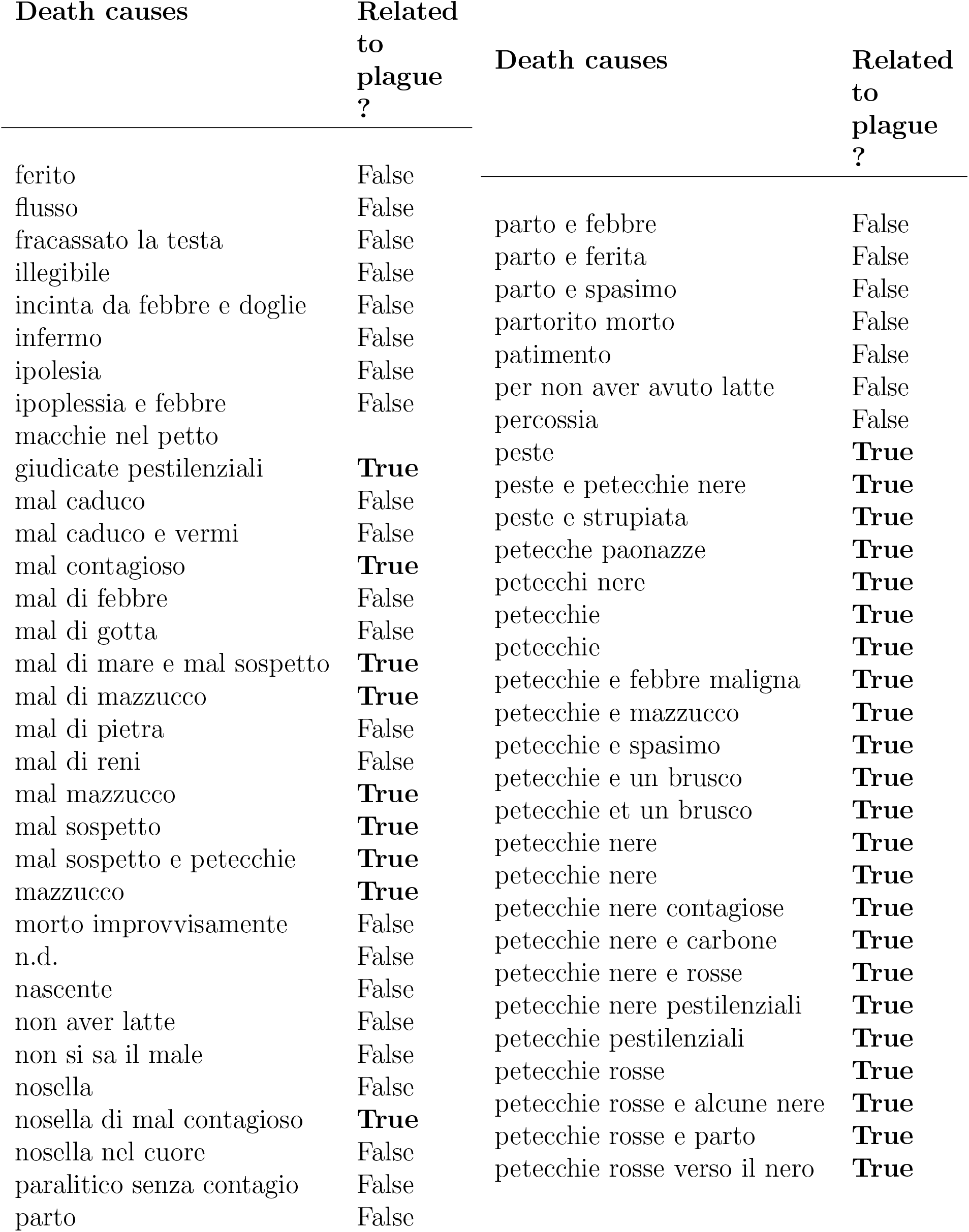
Manual classification of all 156 death causes as reported in necrologies, as associated to plague or not – part 2.

**Supplementary Table SI3:** Manual classification of all 156 death causes as reported in necrologies, as associated to plague or not – part 3.

| Death causes | Related<br>to<br>plague<br>? |
| --- | --- |
| punta | False |
| rogna | False |
| sconosciuta | False |
| sempre infermo | False |
| senza peste esterna | False |
| senza sospetto | False |
| spasimo | False |
| spasimo e infermitá | False |
| spasimo e mazzucco | **True** |
| spasimo e petecchie nere | **True** |
| spasimo e sturioli | False |
| spasimo e vecchiezza | False |
| spasimo e vermi | False |
| stroppiata su la palada | False |
| strupiata | False |
| tumore | False |
| tumore alla gola | False |
| un carbon | **True** |
| variole | False |
| variole e sturioli | False |
| vecchiezza | False |
| vecchiezza e febbre | False |
| vermi | False |
| vermi e petecchie | **True** |
| vermi e spasimo | False |

## Footnotes

1 For more details, find here the description of possible metrics: scipy.spatial.distance.pdist.

## References

[1] Guido Alfani and Tommy E. Murphy. Plague and Lethal Epidemics in the Pre-Industrial World. The Journal of Economic History, 77(01):314–343, March 2017.

[2] M. J. Keeling and C. A. Gilligan. Metapopulation dynamics of bubonic plague. Nature, 407(6806):903–906, 2000.

[3] Michel Drancourt, Linda Houhamdi, and Didier Raoult. Yersinia pestis as a telluric, human ectoparasite-borne organism. The Lancet infectious diseases, 6(4):234–241, 2006.

[4] Anne Karin Hufthammer and Lars Walløe. Rats cannot have been intermediate hosts for Yersinia pestis during medieval plague epidemics in Northern Europe. Journal of Archaeological Science, 40(4):1752–1759, April 2013.

[5] Katharine R. Dean, Fabienne Krauer, Lars Walløe, Ole Christian Lingjærde, Barbara Bramanti, Nils Chr. Stenseth, and Boris V. Schmid. Human ectoparasites and the spread of plague in Europe during the Second Pandemic. Proceedings of the National Academy of Sciences, page 201715640, January 2018.

[6] Mark R. Welford and Brian H. Bossak. Validation of Inverse Seasonal Peak Mortality in Medieval Plagues, Including the Black Death, in Comparison to Modern Yersinia pestis-Variant Diseases. PLoS ONE, 4(12):e8401, 2009.

[7] Boris V. Schmid, Ulf Büntgen, W. Ryan Easterday, Christian Ginzler, Lars Walløe, Barbara Bramanti, and Nils Chr. Stenseth. Climate-driven introduction of the Black Death and successive plague reintroductions into Europe. Proceedings of the National Academy of Sciences, 112(10):3020–3025, March 2015.

[8] G. Christakos, R. A. Olea, and H. L. Yu. Recent results on the spatiotemporal modelling and comparative analysis of Black Death and bubonic plague epidemics. Public Health, 121(9):700–720, 2007.

[9] Kenneth L. Gage and Michael Y. Kosoy. Natural History of the Plague: Perspectives from More than a Century of Research. Annual Review of Entomology, 50(1):505–528, January 2005.

[10] José M. Gómez and Miguel Verdú. Network theory may explain the vulnerability of medieval human settlements to the Black Death pandemic. Scientific Reports, 7:43467, 2017.

[11] Ricci P. H. Yue, Harry F. Lee, and Connor Y. H. Wu. Trade routes and plague transmission in pre-industrial Europe. Scientific Reports, 7(1):12973, December 2017.

[12] Lilith K. Whittles and Xavier Didelot. Epidemiological analysis of the Eyam plague outbreak of 1665–1666. Proceedings of the Royal Society B: Biological Sciences, 283(1830):20160618, May 2016.

[13] Daniel R. Curtis and Joris Roosen. The sex-selective impact of the Black Death and recurring plagues in the Southern Netherlands, 1349-1450. American Journal of Physical Anthropology, 164(2):246–259, October 2017.

[14] Paolo Ulvioni. Il gran castigo di Dio. Carestia ed epidemie a Venezia e nella Ter-raferma 1628-1632. Milano, 1989.

[15] Thi-Nguyen-Ny Tran, Michel Signoli, Luigi Fozzati, Gérard Aboudharam, Didier Raoult, and Michel Drancourt. High Throughput, Multiplexed Pathogen Detection Authenticates Plague Waves in Medieval Venice, Italy. PLoS ONE, 6(3):e16735, March 2011.

[16] Stephen R. Ell. Three days in October of 1630: detailed examination of mortality during an early modern plague epidemic in Venice. Review of Infectious Diseases, 11(1):128–139, 1989.

[17] Alexandra Bamji. Medical Care in Early Modern Venice. Journal of Social History, 49(3):483–509, March 2016.

[18] Guido Alfani and Samuel K Cohn Jr. Nonantola 1630. anatomia di una pestilenza e meccanismi del contagio (con riflessioni a partire dalle epidemie milanesi della prima età moderna). Popolazione e storia, 8(2):99–138, 2007.

[19] Daniele Beltrami. Storia della popolazione di Venezia dalla fine del secolo XVI alla caduta della Repubblica. 1954.

[20] Weiner Gordon M. The Demographic Effects of the Venetian Plagues of 1575-77 and 1630-31. Genus, 26(1/2):41–57, 1970.

[21] Giovanni Favero, Maria Moro, Pierpaolo Spinelli, Francesca Trivellato, and Francesco Vianello. Le anime dei demografi. Fondi per la rilevazione della popolazione di Venezia nei secoli XVI e XVII. Bollettino di demografia storica, 15:23–110, 1991.

[22] Sharon N DeWitte. The effect of sex on risk of mortality during the black death in london, ad 1349–1350. American Journal of Physical Anthropology: The Official Publication of the American Association of Physical Anthropologists, 139(2):222–234, 2009.

[23] Leslie Bradley. The most famous of all english plagues: a detailed analysis of the plague at eyam 1665-6. Local Population Studies, (Supplement 4):63–94, 1977.

[24] Susan Scott and Christopher J Duncan. Biology of plagues: evidence from historical populations. Cambridge University Press, 2001.

[25] Roger Schofield. An anatomy of an epidemic: Colyton november 1645 to november 1646. Local Population Studies, (Supplement 4):95–126, 1977.

[26] Mario Abrate. Popolazione e peste del 1630 [ie un mille seiciento e trenta] a Car-magnola, 1. Centro studi piemontesi, 1972.

[27] Matteo Manfredini, Sergio De Iasio, and Enzo Lucchetti. The plague of 1630 in the territory of parma: Outbreak and effects of a crisis. International Journal of Anthropology, 17(1):41–57, 2002.

[28] Sharon N. DeWitte. Age patterns of mortality during the Black Death in London, A.D. 1349–1350. Journal of Archaeological Science, 37(12):3394–3400, December 2010.

[29] Stefan Monecke, Hannelore Monecke, and Jochen Monecke. Modelling the black death. a historical case study and implications for the epidemiology of bubonic plague. International Journal of Medical Microbiology, 299(8):582–593, 2009.

[30] MJ Keeling and CA Gilligan. Bubonic plague: a metapopulation model of a zoonosis. Proceedings of the Royal Society of London B: Biological Sciences, 267(1458):2219–2230, 2000.

[31] Roger Schofield. The last visitation of the plague in sweden: the case of Bräkne-h oby in 1710–11. The Economic History Review, 69(2):600–626, 2016.

[32] Marcel Salathé and James H Jones. Dynamics and control of diseases in networks with community structure. PLoS computational biology, 6(4):e1000736, 2010.

[33] Sebastian Schnettler. A structured overview of 50 years of small-world research. Social networks, 31(3):165–178, 2009.

[34] Eric Jones, Travis Oliphant, Pearu Peterson, et al. SciPy: Open source scientific tools for Python. https://www.scipy.org/, 2001.

[35] Giulio Rossetti, Letizia Milli, Salvatore Rinzivillo, Alina Ŝirbu, Dino Pedreschi, and Fosca Giannotti. Ndlib: a python library to model and analyze diffusion processes over complex networks. International Journal of Data Science and Analytics, 5(1):61–79, Feb 2018.

[36] Aric A. Hagberg, Daniel A. Schult, and Pieter J. Swart. Exploring network structure, dynamics, and function using networkx. In Gaël Varoquaux, Travis Vaught, and Jarrod Millman, editors, Proceedings of the 7th Python in Science Conference, pages 11 – 15, Pasadena, CA USA, 2008.

